# Research Letter: Incidence of SARS-CoV-2 infection among unvaccinated US adults during the Omicron wave

**DOI:** 10.1101/2022.05.27.22275630

**Authors:** Jennifer L. Alejo, Teresa PY Chiang, Jonathan Mitchell, Aura T. Abedon, Alexa A. Jefferis, William Werbel, Allan B. Massie, Martin A. Makary, Dorry L. Segev

## Abstract

As of 4/20/2022, approximately 23% of the eligible US population was unvaccinated. We studied COVID-19 infections during the Omicron (B.1.1.529) wave in unvaccinated US adults, stratified by pre-Omicron antibody levels. Anti-spike serologic testing was performed prior to the Omicron wave in the United States (9/23/21-11/5/21) and participants were surveilled to determine incident COVID-19. Only 12% of those who entered the wave with antibodies reported a test-confirmed COVID-19 infection, compared to 35% of those without antibodies prior to the Omicron wave. Effectiveness of these anti-RBD antibodies in this unvaccinated population was 67%. Among people with antibodies, titer did not appear to be associated with risk of test-confirmed Omicron infection.

---

As of 4/20/2022, approximately 23% of the eligible US population has not received at least one SARS-CoV-2-vaccine dose.[1] We recently reported that 99% of unvaccinated US adults who had a test-confirmed SARS-CoV-2 infection had detectable anti-RBD antibodies up to 20 months after their positive COVID-19 test.[2] However, whether these antibodies can be used as a clinical marker of protection against new variants is undescribed. To better understand protective correlates, we studied COVID-19 infections during the Omicron (B.1.1.529) wave in unvaccinated US adults, stratified by pre-Omicron antibody levels.

## Methods

Self-described healthy adults who reported never receiving a SARS-CoV-2 vaccine were recruited for this national cohort study between (9/11/2021-10/8/2021).[2] Anti-spike serologic testing was performed prior to the Omicron wave (9/23/21-11/5/21), and categories of antibody titers were created based on reported associations with neutralization.[3-4] Participants completed a follow-up questionnaire (1/19/2022-2/7/2022) about COVID-19 test status and symptoms (since 12/1/2021): tested positive for COVID-19, suspected COVID-19 but never tested positive, or no suspected infection or positive test, and classified symptoms as severe, moderate, mild, or none.

Population characteristics were compared using Fisher’s exact test for categorical variables and Wilcoxon rank-sum test for continuous variables. All analyses were performed using Stata 17.0/SE. The study was approved by the Johns Hopkins institutional review board. Participants provided informed electronic consent.

## Results

Of 843 unvaccinated adults with anti-RBD measured within two months preceding the Omicron wave, 566 (67%) completed the follow-up survey between 1/19/2022-2/7/2022. 20 reported interval COVID-19 vaccination and were excluded. The median (interquartile range, IQR) age was 48 (37-59) years, 295 (54%) were women, 453 (83%) were white, 203 (37%) reported having a test-confirmed COVID-19 diagnosis before the Omicron wave, and 328 (60%) had anti-RBD antibodies before the Omicron wave. 446 (82%) reported no regular public mask use since December 1, 2021. Participants with and without anti-RBD antibodies were similar with respect to age, sex, race, ethnicity, and mask use (Table).

**Table.**
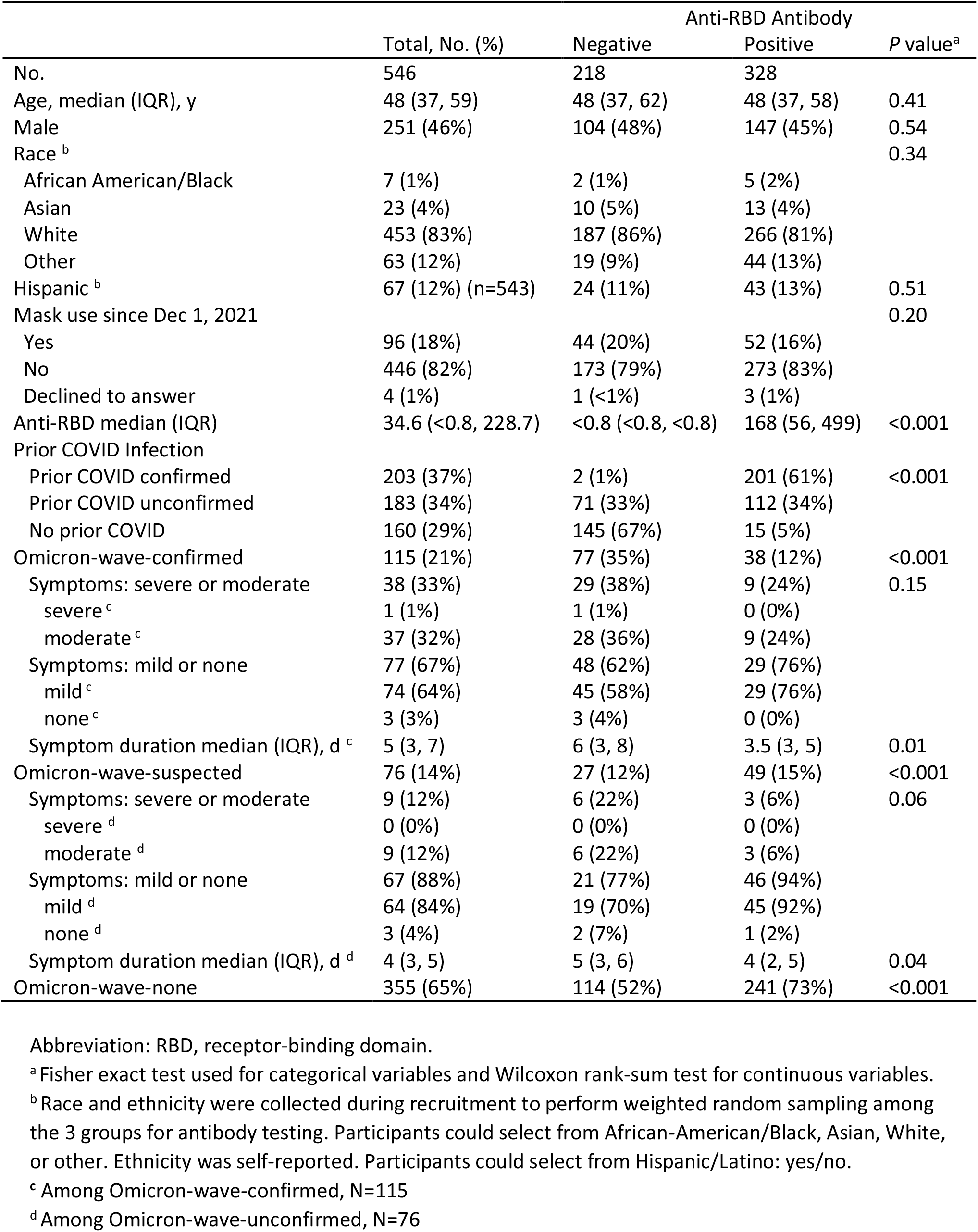
Participant demographics and clinical characteristics, by pre-Omicron anti-RBD level

|  | Total, No. (%) | Anti-RBD Antibody |  | P value<sup>a</sup> |
| --- | --- | --- | --- | --- |
|  |  | Negative | Positive |  |
| No. | 546 | 218 | 328 |  |
| Age, median (IQR), y | 48 (37, 59) | 48 (37, 62) | 48 (37, 58) | 0.41 |
| Male | 251 (46%) | 104 (48%) | 147 (45%) | 0.54 |
| Race<sup>b</sup> |  |  |  | 0.34 |
| African American/Black | 7 (1%) | 2 (1%) | 5 (2%) |  |
| Asian | 23 (4%) | 10 (5%) | 13 (4%) |  |
| White | 453 (83%) | 187 (86%) | 266 (81%) |  |
| Other | 63 (12%) | 19 (9%) | 44 (13%) |  |
| Hispanic<sup>b</sup> | 67 (12%) (n=543) | 24 (11%) | 43 (13%) | 0.51 |
| Mask use since Dec 1, 2021 |  |  |  | 0.20 |
| Yes | 96 (18%) | 44 (20%) | 52 (16%) |  |
| No | 446 (82%) | 173 (79%) | 273 (83%) |  |
| Declined to answer | 4 (1%) | 1 (<1%) | 3 (1%) |  |
| Anti-RBD median (IQR) | 34.6 (<0.8, 228.7) | <0.8 (<0.8, <0.8) | 168 (56, 499) | <0.001 |
| Prior COVID Infection |  |  |  |  |
| Prior COVID confirmed | 203 (37%) | 2 (1%) | 201 (61%) | <0.001 |
| Prior COVID unconfirmed | 183 (34%) | 71 (33%) | 112 (34%) |  |
| No prior COVID | 160 (29%) | 145 (67%) | 15 (5%) |  |
| Omicron-wave-confirmed | 115 (21%) | 77 (35%) | 38 (12%) | <0.001 |
| Symptoms: severe or moderate | 38 (33%) | 29 (38%) | 9 (24%) | 0.15 |
| severe<sup>c</sup> | 1 (1%) | 1 (1%) | 0 (0%) |  |
| moderate<sup>c</sup> | 37 (32%) | 28 (36%) | 9 (24%) |  |
| Symptoms: mild or none | 77 (67%) | 48 (62%) | 29 (76%) |  |
| mild<sup>c</sup> | 74 (64%) | 45 (58%) | 29 (76%) |  |
| none<sup>c</sup> | 3 (3%) | 3 (4%) | 0 (0%) |  |
| Symptom duration median (IQR), d<sup>c</sup> | 5 (3, 7) | 6 (3, 8) | 3.5 (3, 5) | 0.01 |
| Omicron-wave-suspected | 76 (14%) | 27 (12%) | 49 (15%) | <0.001 |
| Symptoms: severe or moderate | 9 (12%) | 6 (22%) | 3 (6%) | 0.06 |
| severe<sup>d</sup> | 0 (0%) | 0 (0%) | 0 (0%) |  |
| moderate<sup>d</sup> | 9 (12%) | 6 (22%) | 3 (6%) |  |
| Symptoms: mild or none | 67 (88%) | 21 (77%) | 46 (94%) |  |
| mild<sup>d</sup> | 64 (84%) | 19 (70%) | 45 (92%) |  |
| none<sup>d</sup> | 3 (4%) | 2 (7%) | 1 (2%) |  |
| Symptom duration median (IQR), d<sup>d</sup> | 4 (3, 5) | 5 (3, 6) | 4 (2, 5) | 0.04 |
| Omicron-wave-none | 355 (65%) | 114 (52%) | 241 (73%) | <0.001 |

Abbreviation: RBD, receptor-binding domain.
<sup>a</sup> Fisher exact test used for categorical variables and Wilcoxon rank-sum test for continuous variables.
<sup>b</sup> Race and ethnicity were collected during recruitment to perform weighted random sampling among the 3 groups for antibody testing. Participants could select from African-American/Black, Asian, White, or other. Ethnicity was self-reported. Participants could select from Hispanic/Latino: yes/no.
<sup>c</sup> Among Omicron-wave-confirmed, N=115
<sup>d</sup> Among Omicron-wave-unconfirmed, N=76

35% of unvaccinated individuals without preexisting antibodies (anti-RBD<0.8 U/mL, n=218) reported test-confirmed COVID-19; an additional 12% reported suspected/unconfirmed COVID-19 during the Omicron wave. In contrast, 12% of unvaccinated individuals with preexisting antibodies (anti-RBD>0.8 U/mL, n=328) reported test-confirmed COVID-19 and an additional 15% reported suspected/unconfirmed COVID-19 during the Omicron wave. Among those with anti-RBD 0.8-1000 U/mL (n=284), 12%/16% reported confirmed/suspected COVID-19, and among those with anti-RBD≥1000 U/mL (n=44), 9%/7% reported confirmed/suspected COVID-19 (Supplementary Figure).

**Figure.**
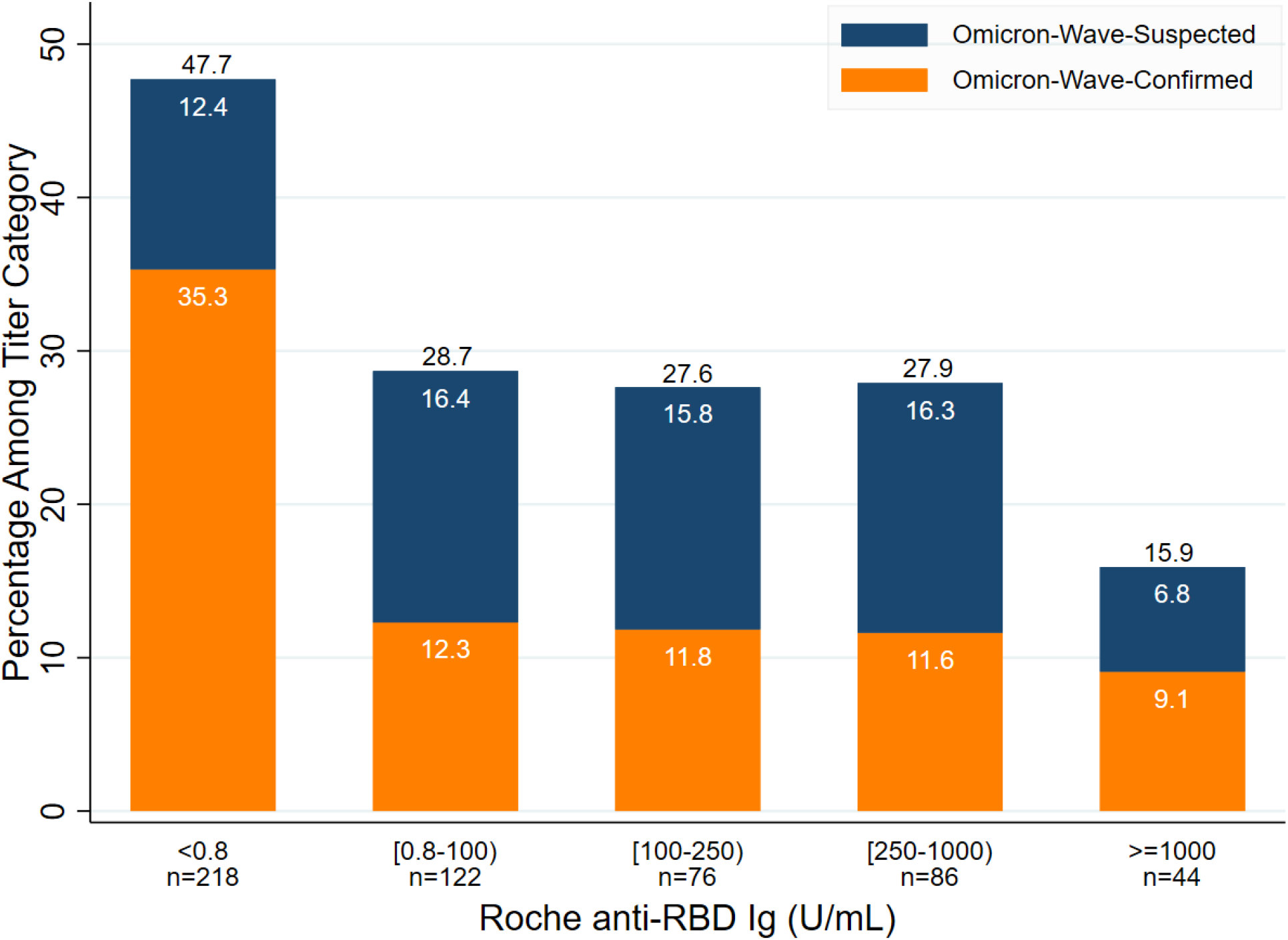
Distribution of self-reported COVID-19 infections after December 1, 2021 among unvaccinated US Adults, stratified by pre-Omicron wave anti-RBD antibody level.

Having antibodies was 67% effective against reporting test-confirmed COVID-19(35% vs 12% p<0.001). Among those with test-confirmed COVID-19 who answered symptom questions (n=115), those with antibodies reported shorter symptom duration than their antibody-negative counterparts (median[IQR] 3.5 [3, 5] vs. 6 [3, 8] days, p=0.013). There was a higher proportion of antibody-negative participants (vs. positive) in the COVID-19-confirmed (38% vs 24%, p=0.15), and suspected/unconfirmed groups (22% vs. 6%, p=0.06), but neither difference was statistically significant.

## Discussion

In this national longitudinal study of incident COVID-19 among unvaccinated adults during the Omicron wave, only 12% of those who entered the wave with anti-RBD antibodies reported a test-confirmed COVID-19 infection, compared to 35% of those without antibodies prior to the Omicron wave. Effectiveness of these anti-RBD antibodies, presumably derived from previous infection, in this unvaccinated population was 67%. Recently, reduced vaccine efficacy against Omicron has been described among 4x-vaccinated Israelis, and the additional protection of a 4^th^ dose peaked four weeks post-fourth dose and waned in later weeks.[4-5] Our results add valuable information to the discussion of vaccine versus infection-derived immune protection against Omicron.[6-7]

Study limitations include lack of information about direct neutralization against Omicron (though anti-RBD correlation with neutralization is described), lack of viral sequencing (though follow-up occurred when Omicron became the dominant strain in the US), self-reported COVID-19 test results, limited availability of COVID-19 testing during the follow-up period which could lead to underreporting of COVID-confirmed cases, and survivor bias.[3]

In conclusion, the presence of anti-RBD antibodies in an unvaccinated healthy adult (natural immunity) was associated with 23% decreased relative risk for COVID-19 reinfection and shorted symptom duration versus those without pre-existing anti-RBD antibodies during the Omicron wave. Among people with antibodies, titer did not appear to be associated with risk of test-confirmed Omicron infection, although our sample size for those ≥1000 U/mL may have been inadequate to detect such a difference in that range. It is important to note that while disease severity for hospitalized Omicron patients was somewhat lower for Omicron versus other variants, patients hospitalized with COVID-19 remain at substantial risk of critical illness and death. [8] Our findings shed some light on COVID protection among the unvaccinated-immune.

## Data Availability

Data produced in the present study are not available.

## Acknowledgement

We acknowledge the following individuals for their assistance with this study, none of whom were compensated for their contributions: Jake D. Kim, BS, and Carolyn N. Sidoti, BS, for data collection and study coordination (Department of Surgery, Johns Hopkins School of Medicine); Daniel S. Warren, PhD (Department of Surgery, Johns Hopkins School of Medicine), Amy Chang, MD, (Department of Surgery, Johns Hopkins School of Medicine) and Macey L. Levan, JD, PhD (Departments of Acute and Chronic Care, Johns Hopkins School of Nursing, and Surgery, Johns Hopkins School of Medicine) for administrative and scientific support.

## Notes

**Conflict of Interest:** DL Segev has the following financial disclosures: consulting and speaking honoraria from Sanofi, Novartis, CLS Behring, Jazz Pharmaceuticals, Veloxis, Mallinckrodt, Thermo Fisher Scientific, Astra Zeneca, and Regeneron. The remaining authors of this manuscript have no financial disclosures or conflicts of interest to disclose.

**Funding/Support:** This work was supported by charitable donations from the Ben-Dov family. These authors received salary support from the following grants: T32DK007713 (Dr. Alejo), The ASTS Jonathan P. Fryer Resident Scientist Scholarship (Dr. Mitchell).

### Competing Interest Statement

DL Segev has the following financial disclosures: consulting and speaking honoraria from Sanofi, Novartis, CLS Behring, Jazz Pharmaceuticals, Veloxis, Mallinckrodt, Thermo Fisher Scientific, Astra Zeneca, and Regeneron. Dr. Levan receives consulting honoraria for Takeda Pharmaceuticals. The remaining authors of this manuscript have no financial disclosures or conflicts of interest to disclose.

### Funding Statement

This work was supported by charitable donations from the Ben-Dov family. These authors received salary support from the following grants: T32DK007713 (Dr. Alejo), The ASTS Jonathan P. Fryer Resident Scientist Scholarship (Dr. Mitchell).

### Author Declarations

Ethics committee/IRB of the Johns Hopkins University gave ethical approval for this work.

